# One-Year Outcomes in Acute Strokes with Hyperglycemia in Low Resource Settings (SHAPE): A Prospective Cohort Study Protocol

**DOI:** 10.1101/2024.12.29.24319757

**Authors:** Bibek Rajbhandari, Yogendra Man Shakya, Ramesh Kumar Maharjan, Dipak Malla, Paras Thapa, Prakash Regmi, Bikal Shrestha, Sumit Shahi

**Affiliations:** Department of General Practice and Emergency Medicine, Maharajgung Medical Campus, Institute of Medicine, Kathmandu, Nepal; Department of Emergency Medicine, Maharajgung Medical Campus, Institute of Medicine, Kathmandu, Nepal; Department of Endocrinology, Bir Hospital, Kathmandu, Nepal; Department of Radiology, Kathmandu Medical College, Kathmandu, Nepal; Department of Neurosurgery, Maharajgung Medical Campus, Institute of Medicine, Kathmandu, Nepal; Department of Community Medicine, Nepalese Army Institute of Health sciences; Department of Neuromedicine, Maharajgung Medical Campus, Institute of Medicine, Kathmandu, Nepal

**Keywords:** *Stroke*, *Hyperglycemia*, *Mortality*, *Cohort study*, *Glycemic levels*

## Abstract

**Background:** Stroke is a significant global health issue, serving as a leading cause of death and disability. The burden of this condition is especially severe in low- and middle-income countries, where the majority of stroke-related fatalities occur. Hyperglycemia, a common metabolic disturbance observed in acute stroke patients, is known to worsen clinical outcomes, contributing to higher rates of mortality and morbidity. This study aims to investigate the relationship between hyperglycemia, morbidity and mortality outcomes in patients with acute stroke, assessing how varying glycemic levels influence short-term (1 month), mid-term (3 months), and long-term (1 year) mortality rates.

**Methods:** The SHAPE study is a prospective cohort study conducted at the Emergency Department of Tribhuvan University Teaching Hospital (TUTH). The study will enroll adults aged over 40 who present with BEFAST-positive symptoms of acute stroke, confirmed through CT/MRI imaging. Participants will be classified into two groups: those exhibiting hyperglycemia (exposed group) and those with normal glycemic levels (unexposed group). Sixty participants will be recruited (30 hyperglycemic and 30 normoglycemic) and followed for one year to assess mortality and functional outcomes using the modified Rankin Scale (mRS). Data collection will involve regular follow-ups through phone calls, clinic visits, and home visits at 1, 3, and 12 months post-stroke. The analysis will include bivariate comparisons for categorical and continuous variables, logistic regression to identify independent predictors of mortality and functional outcomes, and Kaplan-Meier analysis for assessing survival rates.

**Ethical Considerations:** The SHAPE Study was approved by the Ethics Review Committee of the Institute of Medicine (Ref no.. 6-11E2), and written informed consent will be obtained from all participants. Results will be disseminated via a peer-reviewed journal.

**Trial registration number:** ClinicalTrials.gov ID NCT06560983

## INTRODUCTION

Stroke is a major global health issue, ranking as the second leading cause of death and the third leading cause of disability worldwide.^1^ Stroke is responsible for 6.6 million deaths and 143 million disability-adjusted life years (DALYs).^2,3^ The burden is particularly high in low- and middle-income countries (LMICs), where 86% of stroke-related deaths occur.^1^

Among several modifiable risk factors for poor outcomes in acute stroke^4^ , hyperglycemia has emerged as a critical predictor of increased mortality and long-term disability.^5^. Stress-induced hyperglycemia or undiagnosed diabetes during acute stroke^6^ events is known to exacerbate brain injury and impair recovery. Despite advancements in stroke care, hyperglycemia’s impact on patient outcomes remains underexplored, particularly in resource-limited settings where access to timely diagnosis and treatment is constrained.^7^

In Nepal, stroke is a growing public health concern. According to recent data, stroke is the fourth leading cause of death in the country, contributing to a significant portion of non-communicable disease (NCD) mortality.^8^ Local Nepalese doctors and stroke patients’ families founded the Jaya Stroke Foundation, a non-governmental organization. This foundation estimates that 50,000 people per year are afflicted with stroke, with 15,000 people dying from stroke annually.^9^ Limited access to healthcare facilities and diagnostic tools in rural and low-resource settings exacerbates the challenges faced by stroke patients. A systematic review on stroke epidemiology in Nepal revealed an increasing prevalence of ischemic stroke, which often co-occurs with hyperglycemia and other metabolic disturbances.^10^ However, detailed evidence linking hyperglycemia with stroke outcomes in Nepalese populations is lacking, highlighting a critical gap in research and care.^11^ This is especially concerning in settings like the Tribhuvan University Teaching Hospital (TUTH), a significant referral facility in Nepal, where delayed presentations and scarce resources frequently make the management of acute stroke difficult.^12^

Most existing research on hyperglycemia, a modifiable risk factor for poor outcomes in stroke patients, originates from high-income countries with robust healthcare systems. This creates a knowledge gap regarding its impact in LMICs like Nepal, where healthcare challenges are markedly different.^13^ To address this critical gap, this study investigates one-year outcomes in acute stroke patients with hyperglycemia in a low-resource setting in Nepal. This study aims to give important information about the role of hyperglycemia in acute stroke outcomes by looking at the link between glycemic levels and death or morbidity over short-term (1 month), mid-term (3 months), and long-term (1 year) follow-ups. The study will also explore potential factors that may influence the relationship between hyperglycemia and stroke outcomes, such as age, sex, comorbidities and type of stroke. By identifying these factors, healthcare providers can better tailor interventions to improve outcomes for stroke patients with hyperglycemia in similar settings.

## METHODS

### Study design

The SHAPE study is a prospective cohort study designed to examine mortality and morbidity outcomes in hyperglycemic patients with acute stroke. Patients presenting with acute stroke symptoms at the Emergency Department of Tribhuvan University Teaching Hospital (TUTH) will be classified into two cohorts: those with hyperglycemia and those with normal glycemic levels. The participants will be followed over one year with evaluations at one month, three months, and twelve months.

### Study site

The study will be conducted in the Emergency Department of TUTH, one of the largest tertiary care hospitals in Nepal. TUTH handles a high volume of diverse patients, including those presenting with acute stroke from all over Nepal. According to previous data from the TUTH Emergency Department, the prevalence of stroke was 2.96% [95% CI: 2.86-3.10], with ischemic stroke being the most common type, accounting for 76.34% [95% CI: 73.52-79.06].^14^ This setting provides an ideal environment for recruiting and studying stroke patients, particularly in the context of resource-limited settings.

### Study population

The study population consists of a cohort of acute stroke patients, specifically focusing on individuals presenting with acute stroke in the Emergency Room.

### Inclusion Criteria

1. Individuals exhibiting BEFAST-positive symptoms - meeting at least one of the criteria within 24 hours of symptom onset.
2. Confirmation of acute stroke (Ischemic and Hemorrhagic stroke) through radiological imaging.
3. Participants aged over 40 years.

### Exclusion Criteria

1. Patients presenting with hypoglycemia
2. Transient Ischemic Attack (TIA).
3. Subdural hematoma cases.
4. Subarachnoid hemorrhage cases.
5. Diabetic Ketoacidosis.
6. Conditions mimicking stroke symptoms (e.g., sepsis, metabolic derangement, space-occupying lesions, hepatic encephalopathy, uremic encephalopathy).
7. History of baseline bedridden patients.
8. Acute-on-chronic stroke cases.
9. Patients requiring assistance in daily activities.
10. History of trauma preceding the stroke.

In this prospective cohort study, we have established both exposed and unexposed group to facilitate our investigation. The exposed group comprises individuals who meet the criteria for acute stroke and also present with concurrent hyperglycemia. This group will be closely monitored to assess the impact of hyperglycemia on acute stroke outcomes. In contrast, the unexposed group consists of individuals who meet the same criteria for acute stroke but maintain normal glycemic levels.

#### Matching

Matching between the exposed and unexposed groups will be conducted based on age, gender, type of stroke, stroke severity, and treatment received. Stroke severity will be determined using the National Institutes of Health Stroke Scale (NIHSS) for ischemic strokes and the Intracerebral Hemorrhage (ICH) score for hemorrhagic strokes.

#### Sample size

The sample size was calculated using the formula described by Kelsey et. al.^15^ The sample size calculation utilized the following parameters: P1 represents the proportion of unexposed individuals with the outcome or disease, set at 23.9%, while P0 indicates the proportion of exposed individuals with the outcome, which is 68.9%. Both P1 and P0 were taken from a similar study.^16^ A two-tailed confidence interval (CI) of 95% was established, corresponding to Z l1/2 of 1.96, and a statistical power of 80% was defined, indicated by Z **β** of 0.84. The sample size was calculated using OpenEpi version 3.^17^ Based on the outlined parameters, the maximum computed sample size was determined to be 46. To account for potential loss to follow-up, a 20% adjustment for nonrespondents was applied, resulting in a final calculated sample size of 55. For practicality and rounding, the study’s final sample size was set at 60, with 30 participants allocated to each group.

### Study variables and its measurement

| Variables | Definition | Measurement |
| --- | --- | --- |
| **Outcome** |  |  |
| Mortality | Occurrence of death within specified timeframes (short-term: 1 month, mid-term: 3 months, long-term: 1 year) following acute stroke event. | Number of deaths recorded during the predefined time intervals post-stroke. |
| Morbidity | Functional disability or impairment in daily activities resulting from stroke, measured using the modified Rankin Score (MRS).<sup>18</sup> | mRS Score (1-5); 1 = no significant disability, 5 = severe disability/bedridden. Assessed short-term (1 month), mid-term (3 months), and long-term (1 year). |
| **Exposure** |  |  |
| **Demographic Characteristics** |  |  |
| Age | Age of the participants in years. | Recorded in years. |
| Sex | Biological sex of participants. | Male/Female. |
| Occupation | Employment type or professional status of participants. | Categories or specified type of occupation. |
| Ethnicity | Ethnic background or group identity of participants. | Self-reported ethnic group. |
| Height | Participants' height in centimeters (cm). | Measured in centimeters (cm). |
| Weight | Participants' weight in kilograms (kg). | Measured in kilograms (kg). |
| **Behavioral Characteristics** |  |  |
| Physical Activity | Level of physical activity of participants. | Exercise per minute/weekly |
| Smoking | Tobacco usage by participants. | Yes/No/former |
| Alcohol Consumption | Alcohol usage frequency among participants. | Sometimes/Often/Never. |
| **Clinical History and Signs** |  |  |
| Vital Signs | Physiological parameters of health, including pulse, BP, RR, SPO2, temperature, pupil reaction, and Babinski sign. | Pulse (bpm), BP (mmHg), RR (breaths/min), SPO2 (%), temperature (°C/°F), pupil reaction, Babinski sign. |
| Glasgow Coma Scale (GCS) Score | Level of consciousness and neurological function, based on eye, verbal, and motor responses.<sup>19</sup> | GCS score (3-15); higher scores indicate better function. |
| BE FAST Criteria | Stroke symptom detection criteria.<sup>20</sup> | Balance, Eyes (vision), Face drooping, Arm weakness, Speech difficulty, Time to emergency services. |
| Other Symptoms | Reported symptoms like vomiting, headache, and seizures. | Documented symptom occurrence. |
| History of Diabetes, Hypertension, RHD | Participants' medical history of diabetes, hypertension, and rheumatic heart disease. | Yes/No (self-reported or medically documented). |
| **Laboratory Parameters** |  |  |
| GRBS | Random blood sugar levels of participants. | Measured in mg/dL; Target Blood Sugar (140-180mg/dL) or hyperglycemia (>180 mg/dL).<sup>21</sup> |
| **CT /MRI Scan (Stroke Diagnosis)** | Computed Tomography scan (CT) or Magnetic resonance imaging (MRI) will be performed during ER admission |  |
| Hemorrhagic Stroke | Stroke caused by brain bleeding, measured by Intracerebral Hemorrhage (ICH) score and midline shift. | ICH score<sup>22</sup> (assessing severity), midline shift (recorded from CT scan). |
| Ischemic Stroke | Stroke caused by obstruction of blood supply to the brain, measured by CT/MRI findings. | CT/MRI findings (location, size of the affected area). |
| Stroke Severity Assessment | Assessment of stroke severity using National Institutes of Health Stroke Scale (NIHSS) score. | NIHSS score (neurological deficits assessment).<sup>23</sup> |
| **Treatment** |  |  |
| Thrombolysis | Administration of clot-dissolving medication in ischemic stroke cases. | Documented instances of thrombolysis treatment. |

Figure 1 illustrates the study procedure and the participant recruitment process, which follows the hospital protocol.

**Figure 1.**
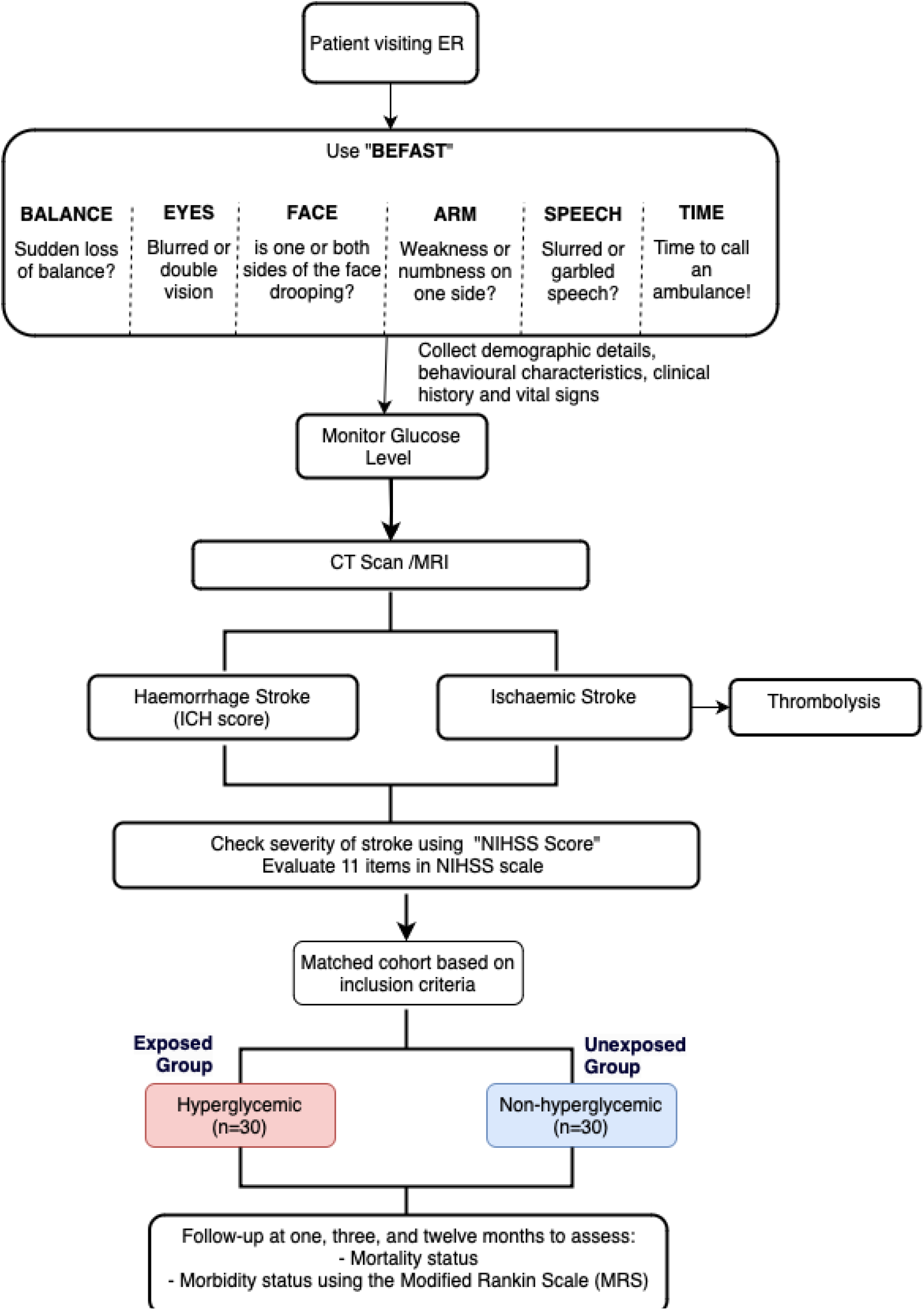
Study procedures

### Assessment: Baseline

Upon arrival at the Emergency Room (ER), patients will be initially assessed using the BEFAST criteria. For those testing positive, demographic details, behavioral characteristics, clinical history, and vital signs will be recorded. Following this, blood glucose levels will be measured using the Glucometer Random Blood Sugar (GBRS) test. Afterward, patients will undergo a CT scan/MRI to classify the type of stroke. Based on the study’s inclusion and exclusion criteria, we will recruit 30 patients with hyperglycemic acute stroke and 30 patients with non-hyperglycemic acute stroke for further follow-up. These participants will be monitored closely at one month, three months, and one year to track mortality and morbidity. All data collected will be systematically entered into the kobotoolbox platform^24^ by trained research assistants to ensure accurate and organized data management.

### Assessment: Follow-Up

To ensure tracking and follow-up of participants in this prospective cohort study, we will establish a structured follow-up system. This system will monitor mortality and morbidity outcomes at one month, three months, and twelve months post-stroke. Below is a detailed follow-up plan:

1. ***Initial Enrollment and Contact Information Collection:*** Upon enrollment in the Emergency Department (ED), detailed contact information, including phone numbers, email addresses, home addresses, and alternative contacts, will be collected, with a primary family contact recorded if the patient is unable to provide details.
2. ***Follow-Up Mechanism:*** We will conduct follow-up assessments at 1, 3, and 12 months post-stroke using phone calls, clinic visits, and home visits (if patients/family members cannot be reached), collecting data on health status, mortality, and functional outcomes through the modified Rankin Scale (mRS). The modified Rankin Scale (MRS)^18^ is a widely used scale for measuring the degree of disability or dependence in individuals who have suffered a stroke or other neurological conditions. It categorizes the level of disability into several distinct categories, which are as follows:

● 0: No symptoms at all.
● 1: No significant disability. Able to carry out all usual activities, despite some symptoms.
● 2: Slight disability. Able to look after own affairs without assistance, but unable to carry out all previous activities.
● 3: Moderate disability. Requires some help, but can walk unassisted.
● 4: Moderate to severe disability. Unable to walk without assistance and unable to attend to own bodily needs without assistance.
● 5: Severe disability. Requires constant nursing care and attention.
● 6: Dead <u>Adverse event management:</u> Given that stroke patients are the participants in our study, various adverse events (AEs) may occur during patient follow-up. We will implement several measures to address these AEs effectively. All AEs will be immediately reported to the principal investigator (BR) for evaluation of their severity and any connection to the study procedures. If needed, suitable medical interventions will be administered, and comprehensive records of each incident will be maintained. Participants who experience AEs will be closely monitored during their follow-up visits. Furthermore, we will keep track of participants’ addresses to establish referral points for immediate action, and research assistants will be trained to recognize and manage AEs.
3. ***Monitoring and tracking system:*** A dedicated team of research assistants will be responsible for follow-up calls, home visits, and scheduling clinic visits. We will utilize a combination of Google Sheets and Google Calendar to facilitate easy data entry and automated reminders. a) Google Sheets: We will maintain a comprehensive database containing patient contact details, follow-up dates, and other relevant information. The database will include columns for Patient ID, contact information, follow-up dates (1, 3, and 12 months), outcome status (Completed/Not Completed), and notes for each follow-up. The data for each patient during every follow-up will be entered using KOBO Toolbox. b) Google Calendar Integration: We will set up reminders for each follow-up in Google Calendar. Follow-up dates will be manually inputted from Google Sheets. These reminders will help ensure that we remain on track for follow-up at 1, 3, and 12 months.
4. ***Data Quality Assurance:*** To ensure data quality, regular weekly monitoring meetings will be conducted to track follow-up progress and prevent patient loss to follow-up. The data management team will verify the completeness and accuracy of data collected from phone calls, clinic visits, and home visits. Additionally, supervisors will routinely review the follow-up data to identify discrepancies and areas for improvement.

### Missing data

We will adopt strategies to handle missing data such as thorough training for research assistants to minimize data collection errors and ensure consistency during follow-ups. We will utilize Kobotoolbox, which includes validation features for each question, to help manage and handle missing data effectively.. In cases of missing data, we will use simple imputation methods, like carrying forward the last observed value, to maintain the integrity of the dataset while still allowing for accurate analysis. Additionally, we will document the reasons for missing data and monitor its impact on the study’s outcomes, enabling us to refine our data collection methods for future research and ensure the robustness of our findings.

### Data Analysis

Descriptive statistics will summarize baseline characteristics, including demographics and clinical history. Comparative analyses, utilizing t-tests for continuous variables and Chi-square tests for categorical variables, will evaluate differences between the two groups. Mortality outcomes will be examined through Kaplan-Meier survival analysis, with a log-rank test used to compare survival between the hyperglycemia and normal glucose level groups. Additionally, a Cox proportional hazards model will assess the relationship between hyperglycemia and mortality while controlling for potential confounders such as age and stroke severity. Functional outcomes will be analyzed using non-parametric tests to evaluate Modified Rankin Scale scores at different follow-up intervals.

### Ethical Considerations

Ethical approval for this study has been obtained from the Institutional Review Board (Ref no. 6- 11E2). Informed written consent will be obtained from all study participants prior to their enrollment. In cases where participants are unable to provide consent, consent will be obtained from a designated patient representative or family member. Prior to obtaining consent, a detailed explanation of the study’s objectives, procedures, potential risks, and benefits will be provided to ensure that participants or their representatives are fully informed. We are committed to maintaining the privacy and confidentiality of all participants throughout the study, adhering to relevant data protection regulations and ethical guidelines. Additionally, all collected data will be securely stored and accessible only to authorized personnel, ensuring that individual identities remain protected.

## DISCUSSION

The SHAPE study protocol presents a timely and critical investigation into the one-year outcomes of hyperglycemia in patients experiencing acute strokes in low-resource settings. With the rising prevalence of diabetes and its established association with poor stroke outcomes, this prospective cohort study addresses a significant gap in understanding how hyperglycemia affects mortality and morbidity rates among stroke patients in Nepal. By focusing on a population that faces distinct challenges in healthcare access and management, the findings from this study could provide valuable insights for improving stroke care for hyperglycemic patients, particularly in low- and middle-income countries (LMICs).

Most of the existing research linking hyperglycemia to adverse stroke outcomes has been conducted in high-income settings,^6,25–27^ leaving a gap in knowledge regarding LMICs where healthcare infrastructure, patient comorbidities, and risk factors may differ. The SHAPE study aims to fill this gap by focusing on hyperglycemia as a modifiable risk factor and its role in stroke mortality. This focus is particularly important in LMICs, where resources for managing both stroke and diabetes are often limited, and targeted interventions could substantially improve clinical outcomes. Notably, this is the first cohort study in Nepal to investigate hyperglycemia in the context of stroke outcomes, building on previous cross-sectional research that only assessed the prevalence of diabetes in acute stroke patients.^10,13,28^ The cohort design allows for a more detailed exploration of hyperglycemia’s role over time.

One of the major strengths of this study is its comparative approach, involving both an exposed group (hyperglycemic patients) and an unexposed group (normoglycemic patients). This comparative design will help clarify the specific impact of hyperglycemia on stroke outcomes. Additionally, the study employs a robust longitudinal design, which facilitates detailed data collection at multiple time points. This approach will provide valuable insights into the temporal relationship between hyperglycemia and mortality, providing a clearer picture of when and how hyperglycemia exacerbates stroke outcomes. These insights are essential for developing more effective, time-sensitive management strategies in stroke care.

The inclusion of diverse patient demographics from low-resource settings enhances the study’s generalizability, making the findings more applicable to real-world clinical environments in LMICs. Since TUTH is one of the largest tertiary hospitals in the country, receiving patients from across Nepal, the SHAPE study will capture a wide range of patients. This makes it well-suited to generate evidence that reflects the realities of stroke care in resource-limited regions, potentially influencing stroke management guidelines and public health initiatives to improve outcomes for hyperglycemic stroke patients.

The SHAPE study addresses a pressing global health issue by investigating the relationship between hyperglycemia and mortality outcomes in acute stroke patients, particularly within low-resource settings. The findings have the potential to inform clinical practice, guide policy decisions, and ultimately improve survival rates and quality of life for individuals affected by stroke in LMICs. By elucidating the impact of glycemic levels on patient outcomes, this study will contribute to the broader discourse on stroke management and highlight the need for effective glycemic control as a critical component of stroke care.

## Data Availability

Not applicable

## Ethics and dissemination

The SHAPE Study has received ethical approval from the Ethics Review Committee of the Institute of Medicine (6-11E2) and written informed consent will be obtained from all participants. All methods will be performed in accordance with the Declaration of Helsinki. Results will be disseminated via a peer-reviewed journal.

## Author Contributions

BR, YMS, and RM were involved in the conceptualization and design of the study. BR, DM, and PT wrote the initial draft of the manuscript. BR, DM, PT, PR, BK, and SS revised the manuscript and provided critical feedback based on their expertise. All authors contributed to the final draft of the manuscript.

## Fundings

This study is funded by the researchers themselves

## Conflict of Interest: **None**

**Patient and public involvement:** Patients and/or the public were not involved in the design, or conduct, or reporting, or dissemination plans of this research.

**Patient consent for publication**: Not required.

## STRENGTHS AND LIMITATIONS OF THIS STUDY

- The study examines cause-and-effect relationships between hyperglycemia and stroke outcomes, providing stronger clinical evidence.
- It addresses a key knowledge gap by studying stroke outcomes in Nepal, a low-income country, providing insights for similar settings.
- Data collection methods such as: self-reported health status or outcomes, could introduce bias if participants are not consistently monitored or evaluated objectively.
- This study is a single-centre study, which restricts external validity

## Notes

### Competing Interest Statement

The authors have declared no competing interest.

### Clinical Protocols

https://clinicaltrials.gov/study/NCT06560983?cond=NCT06560983&rank=1

### Funding Statement

No funding

### Author Declarations

The SHAPE Study was approved by the Ethics Review Committee of the Institute of Medicine (Ref no.. 6-11E2), and written informed consent will be obtained from all participants. Results will be disseminated via a peer-reviewed journal.

## REFERENCES

1. Feigin VL, Owolabi MO, Abd-Allah F, Akinyemi RO, Bhattacharjee NV, Brainin M, et al. Pragmatic solutions to reduce the global burden of stroke: a World Stroke Organization–Lancet Neurology Commission. Lancet Neurol. 2023 Dec 1;22(12):1160–206.

2. Global, regional, and national burden of stroke and its risk factors, 1990-2019: a systematic analysis for the Global Burden of Disease Study 2019. Lancet Neurol [Internet]. 2021 Oct [cited 2024 Oct 17];20(10). Available from: https://pubmed.ncbi.nlm.nih.gov/34487721/

3. Global burden of 369 diseases and injuries in 204 countries and territories, 1990-2019: a systematic analysis for the Global Burden of Disease Study 2019. Lancet [Internet]. 2020 Oct 17 [cited 2024 Oct 17];396(10258). Available from: https://pubmed.ncbi.nlm.nih.gov/33069326/

4. Li S, Wang Y, Wang W, Zhang Q, Wang A, Zhao X. Stress hyperglycemia is predictive of clinical outcomes in patients with spontaneous intracerebral hemorrhage. BMC Neurol. 2022 Jun 27;22(1):236.

5. Middleton S, McElduff P, Ward J, Grimshaw JM, Dale S, D’Este C, et al. Implementation of evidence-based treatment protocols to manage fever, hyperglycaemia, and swallowing dysfunction in acute stroke (QASC): a cluster randomised controlled trial. Lancet. 2011 Nov 12;378(9804):1699–706.

6. Muscari A, Falcone R, Recinella G, Faccioli L, Forti P, Trossello MP, et al. Prognostic significance of diabetes and stress hyperglycemia in acute stroke patients. Diabetol Metab Syndr. 2022 Aug 29;14:126.

7. Tunkl C, Paudel R, Bajaj S, Thapa L, Tunkl P, Chandra A, et al. Implementing stroke care in a lower-middle-income country: results and recommendations based on an implementation study within the Nepal Stroke Project. Front Neurol. 2023 Oct 24;14:1272076.

8. Pandit A, Arjyal A, Farrar J, Basnyat B. Nepal. Practical Neurology. 2006 Apr 1;6(2):129– 33.

9. Shaik MM, Loo KW, Gan SH. Burden of stroke in Nepal. Int J Stroke. 2012 Aug;7(6):517– 20.

10. Paudel R, Tunkl C, Shrestha S, Subedi RC, Adhikari A, Thapa L, et al. Stroke epidemiology and outcomes of stroke patients in Nepal: a systematic review and meta-analysis. BMC Neurol. 2023 Sep 25;23(1):1–12.

11. Bajracharya S, Shrestha BD, Shrestha S, Gorkhaly MP. Clinical Profile of Stroke in Relation to Glycemic Status of the Patients. Medphoenix. 2022 Feb 16;6(2):7–11.

12. Aryal M, Niraula S, Shahukhal R, Mainali NR, Sigdel S, Giri S, Pandey S, Shakya YL. Management of stroke in emergency department in relation to blood pressure, blood sugar and use of anti-thrombotic agents. Journal of Institute of Medicine Nepal. 2010 Nov 24;32(1):11–4.

13. Rayamajhi P, Khadka J, Bhattarai P, Bohaju A, Adhikari A, Mandal D. Diabetes Mellitus among Patients with Acute Ischemic Stroke Admitted to the Department of Medicine in a Tertiary Care Centre. JNMA J Nepal Med Assoc. 2024 Jan 31;62(269):17.

14. Rajbhandari B, Shakya YM, Maharjan RK, Aryal SS, Shah NA, Yadav M, et al. Stroke Presentations in Emergency Care of Nepal: A Mixed-Methods Study Exploring Epidemiological Characteristics and Delays in Acute Treatment. 2024 Sep 26 [cited 2024 Oct 17]; Available from: https://www.researchsquare.com/article/rs-5142547/latest.pdf

15. Methods in Observational Epidemiology [Internet]. Google Books. [cited 2024 Oct 17]. Available from: https://books.google.com/books/about/Methods_in_Observational_Epidemiology.html?id=Xnz6VgL22osC

16. Tshituta JK, Lepira FB, Kajingulu FP, Makulo JRR, Sumaili EK, Akilimali PZ, et al. Prognostic Signification of Admission Hyperglycemia among Acute Stroke Patients in Intensive Care Units in Kinshasa, the Democratic Republic of the Congo. World J Cardiovasc Dis. 2019 Sep 2;9(9):665–80.

17. Sullivan KM, Mir RA, Dean AG. OpenEpi:Sample Size for X-Sectional,Cohort,and Clinical Trials [Internet]. [cited 2024 Oct 17]. Available from: https://www.openepi.com/SampleSize/SSCohort.htm

18. Broderick JP, Adeoye O, Elm J. Evolution of the Modified Rankin Scale and Its Use in Future Stroke Trials. Stroke [Internet]. 2017 Jul [cited 2024 Oct 17];48(7). Available from: https://pubmed.ncbi.nlm.nih.gov/28626052/

19. Jain S, Iverson LM. Glasgow Coma Scale. In: StatPearls [Internet]. StatPearls Publishing; 2023.

20. Aroor S, Singh R, Goldstein LB. BE-FAST (Balance, Eyes, Face, Arm, Speech, Time). Stroke [Internet]. 2017 [cited 2024 Oct 17]; Available from: https://www.ahajournals.org/doi/10.1161/STROKEAHA.116.015169

21. Gaillard T, Miller E. Guidelines for Stroke Survivors With Diabetes Mellitus. Stroke. 2018 Jun;49(6):e215–7.

22. Hemphill JC, Bonovich DC, Besmertis L, Manley GT, Johnston SC. The ICH score: a simple, reliable grading scale for intracerebral hemorrhage. Stroke [Internet]. 2001 Apr [cited 2024 Oct 17];32(4). Available from: https://pubmed.ncbi.nlm.nih.gov/11283388/

23. Appelros P, Terént A. Characteristics of the National Institute of Health Stroke Scale: results from a population-based stroke cohort at baseline and after one year. Cerebrovasc Dis [Internet]. 2004 [cited 2024 Oct 17];17(1). Available from: https://pubmed.ncbi.nlm.nih.gov/14530634/

24. KoboToolbox [Internet]. KoboToolbox. [cited 2024 Oct 17]. Available from: https://www.kobotoolbox.org/

25. Zhao L, Wang L, Lu M, Hu W, Xiu S. Hyperglycemia is associated with poor in-hospital outcome in elderly patients with acute ischemic stroke. Medicine . 2019 Aug 2;98(31):e16723.

26. Shen D, Cai X, Zhu Q, Heizhati M, Hu J, Song S, et al. Increased stress hyperglycemia ratio at hospital admission in stroke patients are associated with increased in-hospital mortality and length of stay. Diabetol Metab Syndr. 2024 Mar 16;16(1):1–9.

27. Levetan CS. Effect of hyperglycemia on stroke outcomes. Endocrine Practice. 2004 Mar 1;10:34–9.

28. Thapa L, Shrestha S, Kandu R, Ghimire MR, Ghimire S, Chaudhary NK, Pahari B, Bhattarai S, Kharel G, Paudel R, Jalan P, Chandra A, Phuyal S, Adhikari B, Aryal N, Kurmi OP. Prevalence of Stroke and Stroke Risk Factors in a South-Western Community of Nepal. J Stroke Cerebrovasc Dis. 2021 May;30(5):105716.

